# Tobacco-Associated Disease Claims and ICD-10 F17 Tobacco-Dependence Coding Among Psychiatric Patients in Indonesia’s National Health Insurance Dataset: A Retrospective Claims-Based Observational Study, 2015–2023

**DOI:** 10.64898/2026.06.25.26356584

**Authors:** Audrey Natalia, Alvin Johan

**Affiliations:** PT Arya Noble, Jakarta, Indonesia; Department of Internal Medicine, Faculty of Medicine, Universitas Indonesia, Jakarta, Indonesia

**Keywords:** tobacco dependence, smoking cessation, F17 coding, health expenditure, administrative claims, Indonesia

## Abstract

**Objectives:** To compare hospital claims and costs for major tobacco-associated diseases with ICD-10 F17 tobacco-dependence coding in a national health-insurance dataset, testing whether the insurer records tobacco addiction or pays for its complications.

**Design:** Retrospective claims-based observational study of routinely collected administrative claims, reported following STROBE and the RECORD extension.

**Setting:** Indonesia’s national health insurance scheme (JKN), which covers more than 90% of the country’s 280 million people and is one of the world’s largest single-payer systems; referral-hospital and primary-care claims, 2015–2023.

**Participants:** A national mental-health claims sample of 54,820 members with at least one ICD-10 mental or behavioural (F-code) diagnosis (weighted to 1,032,022 members) and their 2,074,277 referral-hospital visits.

**Primary and secondary outcome measures:** Primary: verified claim costs (US$) for hospital visits with a primary diagnosis of COPD (J44), trachea/bronchus/lung cancer (C33–C34), ischaemic heart disease (I20–I25) or stroke (I60–I69). Secondary: counts of F17 tobacco-dependence coding and the disease-to-F17 ratio.

**Results:** The four diseases accounted for 13,946 visits among 5,223 patients and US$4.20 million in verified costs (∼6.0% of the sample’s hospital spending; weighted ∼US$74.7 million), of which cardiovascular and cerebrovascular disease formed 95%. F17 appeared in only 51 hospital and 26 primary-care encounters (disease-to-F17 ratio ≈273:1); just 2 of 5,223 disease patients (0.04%) were ever coded F17.

**Conclusions:** The national insurer paid substantially for tobacco-associated morbidity while tobacco dependence was almost never coded. Smoking-related diseases are reimbursed, but tobacco-dependence treatment is not a financed benefit; embedding brief cessation care, reimbursable pharmacotherapy and routine F17 coding into primary care could convert a downstream cost centre into an addiction-care target.

**Strengths and limitations of this study:**

- The study uses a large national health-insurance claims sample (54,820 members; >2 million referral-hospital visits over nine years) with verified payment amounts (converted from Indonesian rupiah to US dollars), applying official sampling weights and a survey-design (stratified) variance estimator.
- Disease groups and tobacco dependence are identified with an explicit, reproducible ICD-10 code list applied to primary and secondary diagnosis fields across hospital and primary-care claims.
- Reporting follows the STROBE statement and the RECORD extension for routinely collected health data, with a transparent claims-selection flow.
- Administrative claims cannot establish that disease was caused by smoking, and contain no patient-level smoking status, pack-years or behavioural data.
- F17 counts reflect coding and reimbursement practice rather than the true prevalence of tobacco dependence; and, as a psychiatric (mental-health) sample, the cohort may under-capture the severity and cost of tobacco-associated disease and cannot be generalised to the wider smoking population.

## Introduction

Indonesia has one of the highest burdens of tobacco use in the world. The 2021 Global Adult Tobacco Survey estimated that 34.5% of adults—approximately 70.2 million people—used tobacco, including 65.5% of men and 3.3% of women.^1^ Modelling of the working-age population has projected hundreds of thousands of excess deaths and very large health and productivity losses attributable to smoking,^2^ and cost-of-illness analyses have estimated the 2019 economic cost of smoking in Indonesia at approximately US$13–29 billion, equivalent to 1.16–2.59% of gross domestic product.^3^ These figures mirror the global pattern, in which smoking-attributable disease consumed an estimated US$422 billion in healthcare spending—about 5.7% of global health expenditure—in 2012 alone.^4^

Since 2014, the national health insurance scheme (Jaminan Kesehatan Nasional, JKN), administered by BPJS Kesehatan, has provided near-universal coverage—now reaching more than 90% of Indonesia’s population of over 280 million people, one of the largest single-payer health systems in the world—and finances most hospital care in the country. The diseases most strongly associated with smoking—chronic obstructive pulmonary disease (COPD), lung cancer, ischaemic heart disease and stroke—are precisely the high-cost conditions that dominate hospital reimbursement. Although cessation support exists in principle, through Ministry of Health quit-line services and selected clinics, tobacco dependence is rarely managed as a reimbursable clinical condition in routine practice, and pharmacotherapy for cessation is not covered by the scheme.

Whether a health system records and treats tobacco dependence, or merely pays for its complications, can be examined directly in its claims data. The ICD-10 category F17 (mental and behavioural disorders due to use of tobacco) is the diagnostic label through which addiction becomes visible to an insurer. International claims studies show that tobacco is heavily under-coded: among United States Medicare beneficiaries, administrative tobacco-use codes rose over time but remained far below survey-based prevalence,^5^ and analyses of Medicaid claims show low rates of tobacco-use-disorder diagnosis and cessation-treatment billing.^6,7^ This under-coding matters most for populations at highest risk. People with mental and behavioural disorders smoke at more than twice the rate of the general population—two to three times in schizophrenia^8^—and bear a disproportionate share of tobacco harm, yet are least likely to receive coverage-dependent dependence treatment.^7^

National-insurer claims have been used elsewhere to quantify the fiscal weight of smoking—for example, South Korea’s National Health Insurance Service attributed US$29.9 billion of medical spending to smoking over 2014–2024, of which cardiometabolic disease accounted for 53% and cancers 35%^9^—and similar cost-of-illness work has been reported for India^10^ and Iran, where cardiovascular disease alone drove roughly three-quarters of smoking-attributable hospital costs.^11^ Routinely collected primary- and secondary-care data have likewise been used to cost the downstream consequences of smoking.^12^ To our knowledge, however, no study has used Indonesia’s national-insurance claims to juxtapose the cost of tobacco-associated disease against the coding of tobacco dependence itself.

We therefore analysed a national mental-health claims sample (the Mental Health Contextual Sample, 2015–2023) to compare referral-hospital claims and verified costs for major tobacco-associated diseases with ICD-10 F17 tobacco-dependence coding, and to quantify the resulting disease-to-F17 ratio. Our specific objectives were: (i) to estimate the volume, patient count and verified cost of hospital care for COPD, trachea/bronchus/lung cancer, ischaemic heart disease and stroke; (ii) to count F17-coded encounters across hospital and primary-care claims and in primary and secondary diagnosis fields; and (iii) to express the contrast between the two as a disease-to-F17 ratio.

## Methods

### Reporting guideline and study design

This retrospective claims-based observational study used routinely collected administrative claims and was reported according to the Strengthening the Reporting of Observational Studies in Epidemiology (STROBE) statement^13^ and the REporting of studies Conducted using Observational Routinely collected health Data (RECORD) extension.^14^ A completed STROBE/RECORD checklist is provided as a supplement.

### Setting and data source

Indonesia’s national health insurance scheme (JKN) is a single-payer system administered by BPJS Kesehatan that covers more than 90% of the population—over 270 million people in a country of more than 280 million. We used this psychiatric-contextual sample deliberately: people with mental and behavioural disorders smoke at two to three times the rate of the general population and bear a disproportionate share of tobacco harm, yet are least likely to receive cessation treatment,^7,8^ making them a sentinel population in which to test whether the insurer records dependence or only pays for its complications. We analysed the Mental Health Contextual Sample (Sampel Kontekstual Kesehatan Mental), a de-identified, weighted national sample of members with mental-health service use, covering service dates from 2015 to 2023.^15^

The dataset comprises linked files for membership (demographics, sampling weight and vital status), referral-hospital claims, hospital secondary diagnoses, and primary-care claims (capitation and non-capitation). Files were linked at member level by an anonymised member identifier and at encounter level by a visit identifier.

### Participants and population selection

The sample comprised 54,820 members, each carrying a sampling weight calibrated to the national mental-health population. All 54,820 members had at least one recorded ICD-10 mental or behavioural (F-code) diagnosis across hospital or primary-care claims; the weighted population represented was 1,032,022 members. A member was flagged as an F-code case when any diagnosis field carried an ICD-10 code in the range F00–F99 in any linked file (hospital primary diagnosis, hospital secondary diagnosis, or primary-care diagnosis); no minimum number of F-coded encounters was required. We placed no age restriction, reflecting the all-age composition of the sample. The analysis of disease cost and F17 coding used the 2,074,277 referral-hospital claim records linked to these members, supplemented by 1,743,537 primary-care capitation and 2,486 non-capitation records for the ascertainment of F17 coding in primary care.

### Variables and code lists (RECORD 6.1, 7.1)

Tobacco-associated disease was defined a priori using the primary-diagnosis ICD-10 field of each hospital claim (three-character code), grouped as: COPD (J44); trachea, bronchus and lung cancer (C33–C34); ischaemic heart disease (I20–I25); and stroke/cerebrovascular disease (I60–I69).^16^ Tobacco dependence was defined as ICD-10 F17 (mental and behavioural disorders due to use of tobacco), ascertained from the hospital primary and secondary diagnoses and the primary-care diagnoses (capitation and non-capitation). The full code list is provided as a supplement. The ICD-10 codes were used as recorded by providers and were not independently validated against medical records within this dataset.

### Outcomes and cost measurement

The primary outcome was the verified claim cost—the amount payable after the insurer’s verification, which we used in preference to the provider-billed amount. All costs were converted from Indonesian rupiah to US dollars (see below). Each referral-hospital record represents one verified hospital visit. Visits were counted per record; patients were counted as unique members; and costs were summed within disease groups. The secondary outcome comprised counts of F17-coded encounters (primary and secondary fields, hospital and primary care) and unique F17-coded patients, and the disease-to-F17 ratio (tobacco-associated disease visits divided by F17 encounters).

### Quantitative variables, weighting and statistical methods

Costs are reported in US dollars (US$), converted from Indonesian rupiah (IDR) at the 2015–2023 average official exchange rate of IDR 14,200 per US$1.^17^ We applied a single period-average rate to all years so that reported changes reflect health-care activity rather than exchange-rate movement; this single-rate simplification is noted as a limitation. Costs are presented both as directly observed sample totals and as population-weighted projections obtained by applying the member sampling weight to each record. Weighted member and patient counts were obtained by summing weights over unique members; weighted visit counts by summing weights over records; and weighted costs by summing the product of verified cost and weight. For member-level proportions we estimated 95% confidence intervals (CIs) using a survey (Taylor-linearised) design with the member as the primary sampling unit and district of residence as the stratum, matching the sampling design declared by the insurer. Continuous age (computed at 2023 from date of birth) is summarised as median and interquartile range (IQR). All reported estimates are descriptive and unadjusted; because the study quantifies coded and reimbursed activity rather than testing an exposure–outcome association, no adjustment for confounders was undertaken. Analyses were performed in Python 3 (pandas, NumPy); the analysis code is available as a supplement. To test the robustness of the disease-to-F17 contrast to our coding choices, we pre-specified three sensitivity analyses: (i) ascertaining tobacco-associated disease from any diagnosis field (hospital primary or secondary) rather than the primary-diagnosis field alone; (ii) broadening the COPD definition from J44 to J40–J44; and (iii) excluding claims with missing or non-positive verified cost.

### Data access, cleaning and linkage (RECORD 12.1–12.3)

The investigators analysed the full released Mental Health Contextual Sample—the complete weighted set of five linked files—rather than an extract, and no further sub-sampling was applied beyond the F-code inclusion criterion above. Data preparation was confined to standardising diagnosis codes (trimming and upper-casing the three-character ICD-10 strings), parsing service dates to calendar year, and treating each referral-hospital record as one verified visit; no records were imputed or deleted during cleaning. Linkage was deterministic: member-level files were joined on the anonymised member identifier and the hospital secondary-diagnosis file on the visit identifier. Linkage was effectively complete—all 2,074,277 referral-hospital, 1,743,537 primary-care capitation and 2,486 non-capitation records carried a member identifier that matched the membership file (100%), all 2,240,698 secondary-diagnosis records linked to a hospital visit (100%), and claim-level and membership sampling weights agreed exactly for every record. Of the 54,820 sampled members, 51,026 (93.1%) had at least one referral-hospital claim during 2015–2023. The data-linkage and claims-selection process, with record counts and linkage quality at each stage, is shown in Supplementary Figure S1.

### Bias, study size and missing data

Because the dataset is a complete enumeration of claims for a fixed, weighted sample, no a priori sample-size calculation was performed; precision is reported through 95% CIs for proportions. The principal sources of bias are inherent to claims data: outcomes reflect coded and reimbursed activity rather than clinical truth, and tobacco dependence that is neither diagnosed nor coded is invisible. We addressed avoidable error by using verified rather than billed costs, by restricting disease ascertainment to the primary-diagnosis field (reducing rule-out and historical codes), and by searching all available diagnosis fields and care settings for F17 to avoid understating dependence coding. As a reflection of data-quality filtering, a primary diagnosis was missing or invalid in only 47 of 2,074,277 referral-hospital records (0.002%); these were retained in denominators but could not enter a disease group. Verified cost was missing in 2 records and non-positive in 3 (together <0.001%), and all service dates parsed successfully. Membership variables used for description (sex, date of birth, ward class, contribution segment and district of residence) were complete, with no missing values, so no records were excluded for missing covariates.

### Patient and public involvement

No patients or members of the public were involved in the design, conduct or reporting of this secondary analysis of de-identified administrative data. The findings will be disseminated to relevant policy stakeholders through the national insurer and professional channels.

### Ethics

The study used a de-identified, governance-controlled secondary dataset released by BPJS Kesehatan and contains no directly identifying information. Analysis of this de-identified sample did not constitute human-subjects research requiring individual consent; use of the data was subject to BPJS data-use governance. No attempt was made to re-identify individuals.

## Results

### Participants

The sample comprised 54,820 members (weighted 1,032,022), all with at least one recorded mental or behavioural (F-code) diagnosis (Table 1). Members were 50.4% male (weighted 52.3%, 95% CI 51.8–52.7); the median age was 36 years (IQR 20–53; weighted median 35, IQR 19–51). Nearly one-quarter (23.2% weighted) were younger than 18 years. The sample was socio-economically skewed toward subsidised and lower-class members: 47.5% were subsidised contribution-assistance beneficiaries and 64.1% were entitled to the lowest hospital ward class. Vital-status records indicated 999 deaths (1.8%) during 2015–2023. These members generated 2,074,277 referral-hospital visits (weighted 40,196,048), with a total verified hospital cost of US$70.6 million. The cohort was dominated by severe mental illness: schizophrenia (F20) was by far the most common psychiatric primary diagnosis (39,233 hospital visits among 10,447 patients) and also the most costly in aggregate (US$6.4 million), followed by other anxiety disorders (F41; 9,749 visits), depressive episode (F32; 7,487 visits), schizoaffective disorder (F25; 4,065 visits) and bipolar disorder (F31; 2,957 visits); developmental disorders (speech and language, F80; pervasive developmental disorders, F84) were common among younger members. The highest median cost per encounter was for delirium (F05; about US$473 per visit), reflecting acute inpatient care. This preponderance of severe mental illness—conditions carrying two- to three-fold higher smoking prevalence—is the context for the tobacco findings that follow. Membership characteristics were complete, with no missing values for sex, age, ward class, contribution segment or district of residence, and a primary diagnosis was missing or invalid in only 47 of 2,074,277 hospital visits (0.002%). The claims-selection flow is shown in Figure 1.

**Figure 1.**
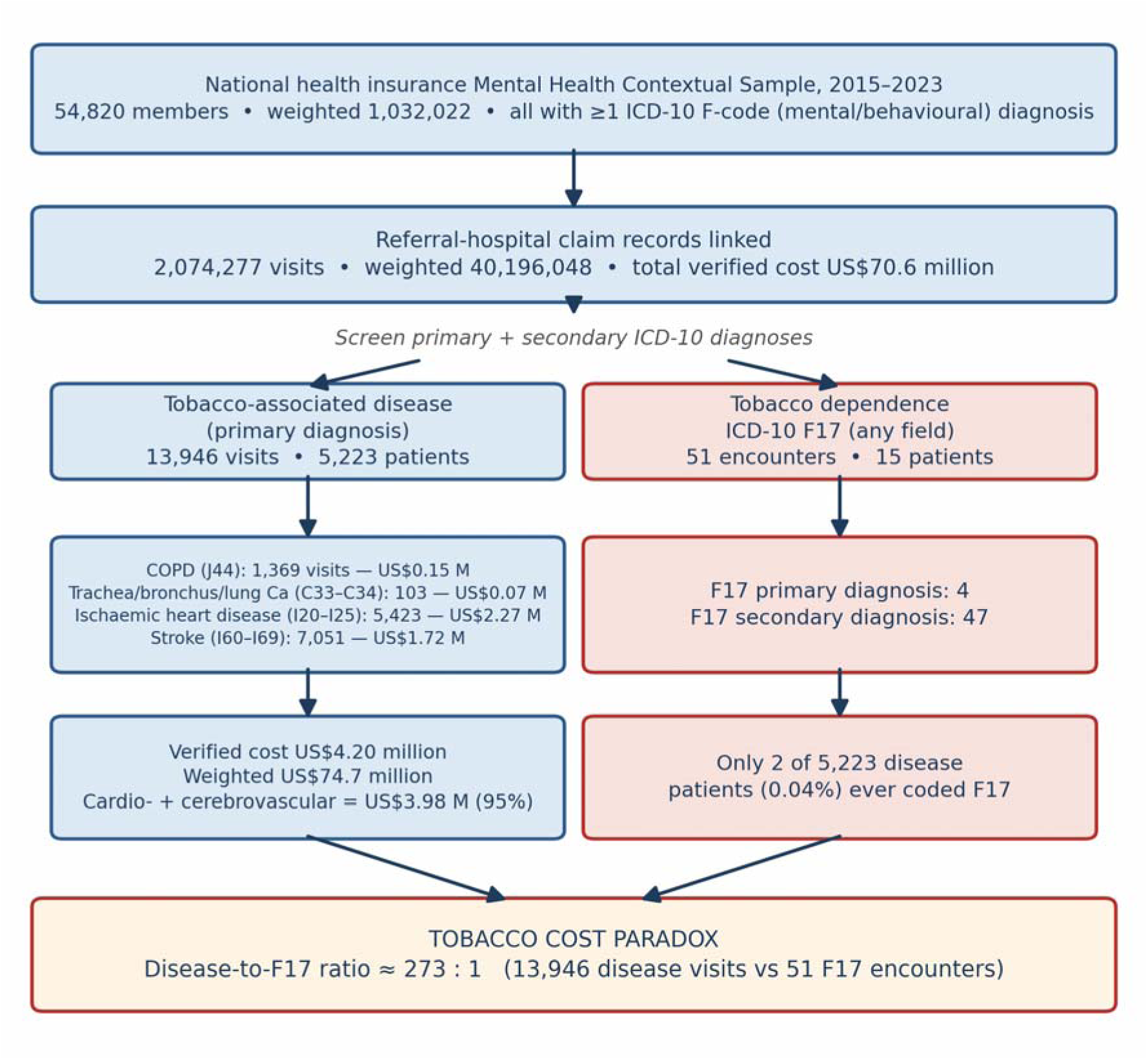
Claims-selection flow and the tobacco cost paradox, national mental-health claims sample 2015–2023. The sample of 54,820 members (weighted 1,032,022) generated 2,074,277 referral-hospital visits; primary-diagnosis screening identified four tobacco-associated disease groups (13,946 visits; 5,223 patients; US$4.20 million verified), against which ICD-10 F17 coding (51 hospital encounters; 15 patients) is contrasted. The full data-linkage and claims-selection flow, with per-stage record counts and linkage quality, is provided as Supplementary Figure S1.

**Table 1.** Characteristics of the national mental-health claims sample, 2015–2023 (N=54,820 members).

| Characteristic | Unweighted n (%) | Weighted % (95% CI) |
| --- | --- | --- |
| Members | 54,820 (100) | 1,032,022 |
| Male sex | 27,620 (50.4) | 52.3 (51.8–52.7) |
| Female sex | 27,200 (49.6) | 47.7 (47.3–48.2) |
| Age, years — median (IQR) | 36 (20–53) | 35 (19–51) |
| Age <18 | — | 23.2 |
| Age 18–29 | — | 18.3 |
| Age 30–44 | — | 23.6 |
| Age 45–59 | — | 20.3 |
| Age ≥60 | — | 14.6 |
| ≥1 mental/behavioural (F) diagnosis | 54,820 (100) | 100 |
| Subsidised beneficiary (PBI) | — | 47.5 |
| Lowest hospital class (class 3) | — | 64.1 |
| ≥1 of four tobacco-associated diseases | 5,223 (9.5) | 8.67 (8.40–8.94) |
| Death recorded 2015–2023 | 999 (1.8) | 17,239 (weighted n) |

CI, confidence interval; IQR, interquartile range; PBI, contribution-assistance/subsidised beneficiaries. Weighted estimates apply the member sampling weight; 95% CIs use a survey design stratified by district of residence.

### Tobacco-associated disease: visits, patients and cost

A primary diagnosis of one of the four tobacco-associated disease groups was recorded in 13,946 hospital visits among 5,223 unique patients (9.5% of members; weighted prevalence 8.67%, 95% CI 8.40–8.94), accounting for US$4.20 million in verified costs—about 6.0% of the sample’s total verified hospital spending—and projecting to approximately 232,339 visits, 89,481 patients and US$74.7 million when weighted to the national mental-health population (Table 2).

**Table 2.** Tobacco-associated disease groups: referral-hospital visits, patients and verified cost (US$), observed and weighted (2015–2023).

| Disease group (ICD-10) | Visits | Patients | Verified cost (US\$ million) | Median cost/visit (US\$) | Weighted visits | Weighted patients | Weighted cost (US\$ million) |
| --- | --- | --- | --- | --- | --- | --- | --- |
| COPD (J44) | 1,369 | 605 | 0.15 | 22 | 20,869 | 9,590 | 2.15 |
| Trachea/bronchus/lung cancer (C33–C34) | 103 | 37 | 0.07 | 473 | 2,025 | 586 | 1.34 |
| Ischaemic heart disease (I20–I25) | 5,423 | 2,294 | 2.27 | 82 | 80,643 | 35,278 | 37.87 |
| Stroke / cerebrovascular (I60–I69) | 7,051 | 2,902 | 1.72 | 22 | 128,802 | 54,059 | 33.39 |
| **Cardio- + cerebrovascular subtotal** | **12,474** | **—** | **3.98** | **—** | **209,445** | **—** | **71.26** |
| **All four groups** | **13,946** | **5,223** | **4.20** | **—** | **232,339** | **89,481** | **74.75** |

Disease groups defined on the primary-diagnosis field. Costs were converted from Indonesian rupiah to US dollars at IDR 14,200 per US\$1 (2015–2023 average official exchange rate). Patient counts do not sum across rows because a patient may have more than one disease; the all-four total is unique patients. Verified cost is the post-verification payable amount. Weighted estimates apply member sampling weights to project to the national mental-health population.

Cardiovascular and cerebrovascular disease dominated. Stroke (I60–I69) accounted for 7,051 visits (US$1.72 million) and ischaemic heart disease (I20–I25) for 5,423 visits (US$2.27 million); together these two groups represented 12,474 visits and US$3.98 million—95% of the four-group cost (weighted US$71.3 million). COPD (J44) contributed 1,369 visits (US$0.15 million) and trachea/bronchus/lung cancer (C33–C34) 103 visits (US$0.07 million) (Figure 2). Cancer was the most expensive per visit (median US$473) and ischaemic heart disease the next (median US$82), whereas COPD and stroke had lower median per-visit costs (US$22 each), consistent with a mix of out-patient follow-up and acute admissions. Member-level weighted prevalence was 5.24% for stroke (95% CI 5.02–5.45), 3.42% for ischaemic heart disease (3.25–3.59), 0.93% for COPD (0.84–1.02) and 0.06% for lung cancer (0.03–0.08).

**Figure 2.**
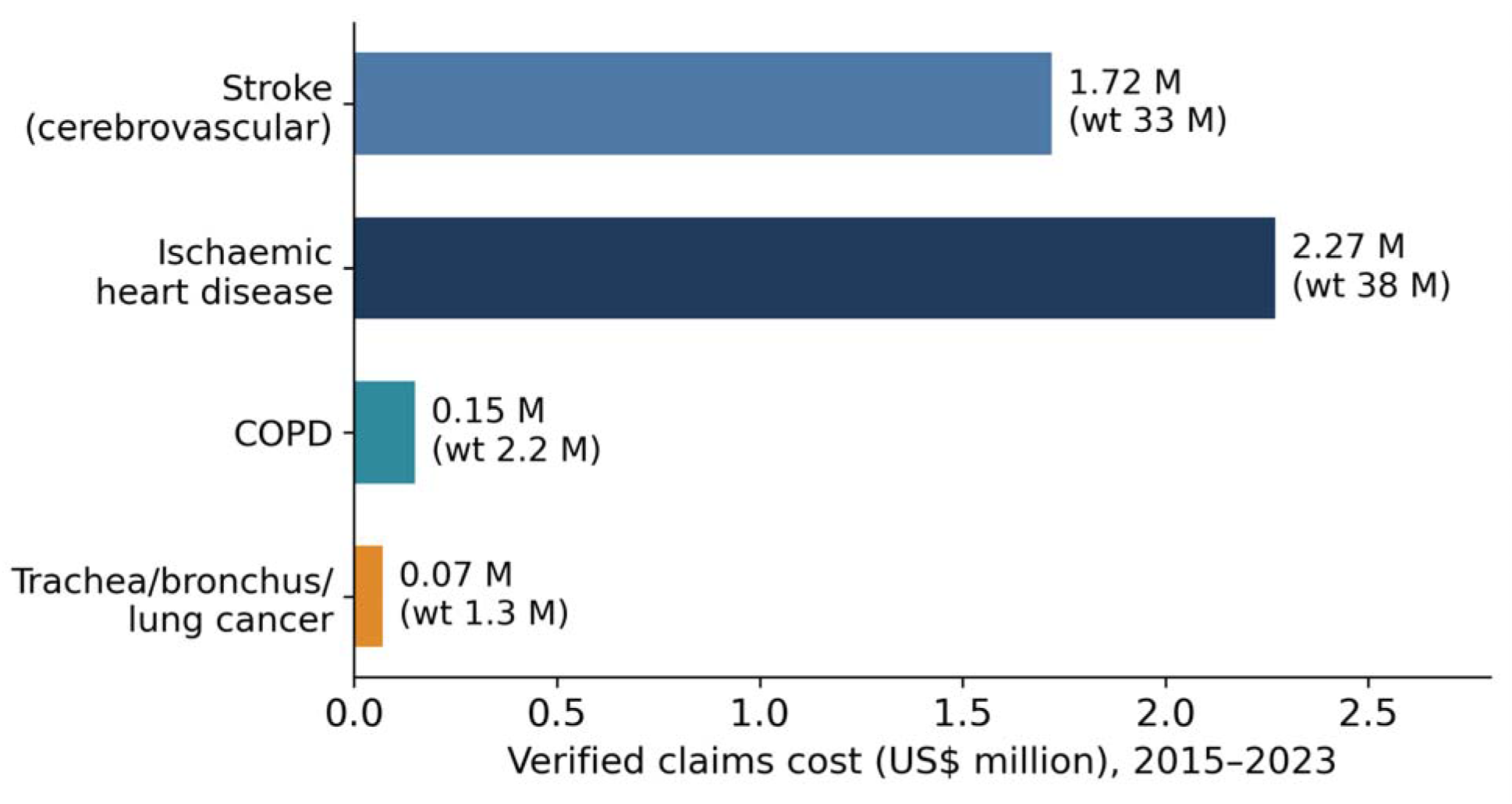
Verified hospital-claims cost by tobacco-associated disease group, 2015–2023 (US$ million; weighted projection in parentheses).

### Tobacco-dependence (F17) coding

Against this burden, ICD-10 F17 tobacco-dependence coding was almost entirely absent (Table 3). Across 2,074,277 hospital visits, F17 appeared as a primary diagnosis in only 4 encounters and as a secondary diagnosis in 47, a total of 51 hospital encounters among just 15 patients. In primary care, F17 was recorded in only 26 capitation visits (17 patients) and no non-capitation visits—77 F17-coded encounters in total across all settings over nine years, in a sample representing more than one million members.

**Table 3.** ICD-10 F17 tobacco-dependence coding and the disease-to-F17 ratio.

| Measure | Value |
| --- | --- |
| F17 as primary diagnosis (hospital) | 4 encounters |
| F17 as secondary diagnosis (hospital) | 47 encounters |
| F17 any field (hospital) — encounters / patients | 51 / 15 |
| F17 in primary care (capitation) — visits / patients | 26 / 17 |
| F17 in primary care (non-capitation) | 0 |
| Total F17-coded encounters, all settings | 77 |
| Tobacco-associated disease visits (hospital) | 13,946 |
| **Disease-to-F17 ratio (hospital encounters)** | **≈273:1** |
| **Disease patients ever coded F17** | **2 of 5,223 (0.04%)** |

F17, mental and behavioural disorders due to use of tobacco. Counts are directly observed (unweighted) sample values over 2015–2023; F17 events are too few for stable population-weighted projection, so weighted estimates are not reported.

The contrast between disease cost and dependence coding defines the central finding. The 13,946 tobacco-associated disease visits outnumbered the 51 hospital F17 encounters by approximately 273 to 1 (Figure 3). Most strikingly, of the 5,223 patients hospitalised with a tobacco-associated disease—the population in whom tobacco dependence is most clinically relevant—only 2 (0.04%) were ever assigned an F17 code in any field or setting.

**Figure 3.**
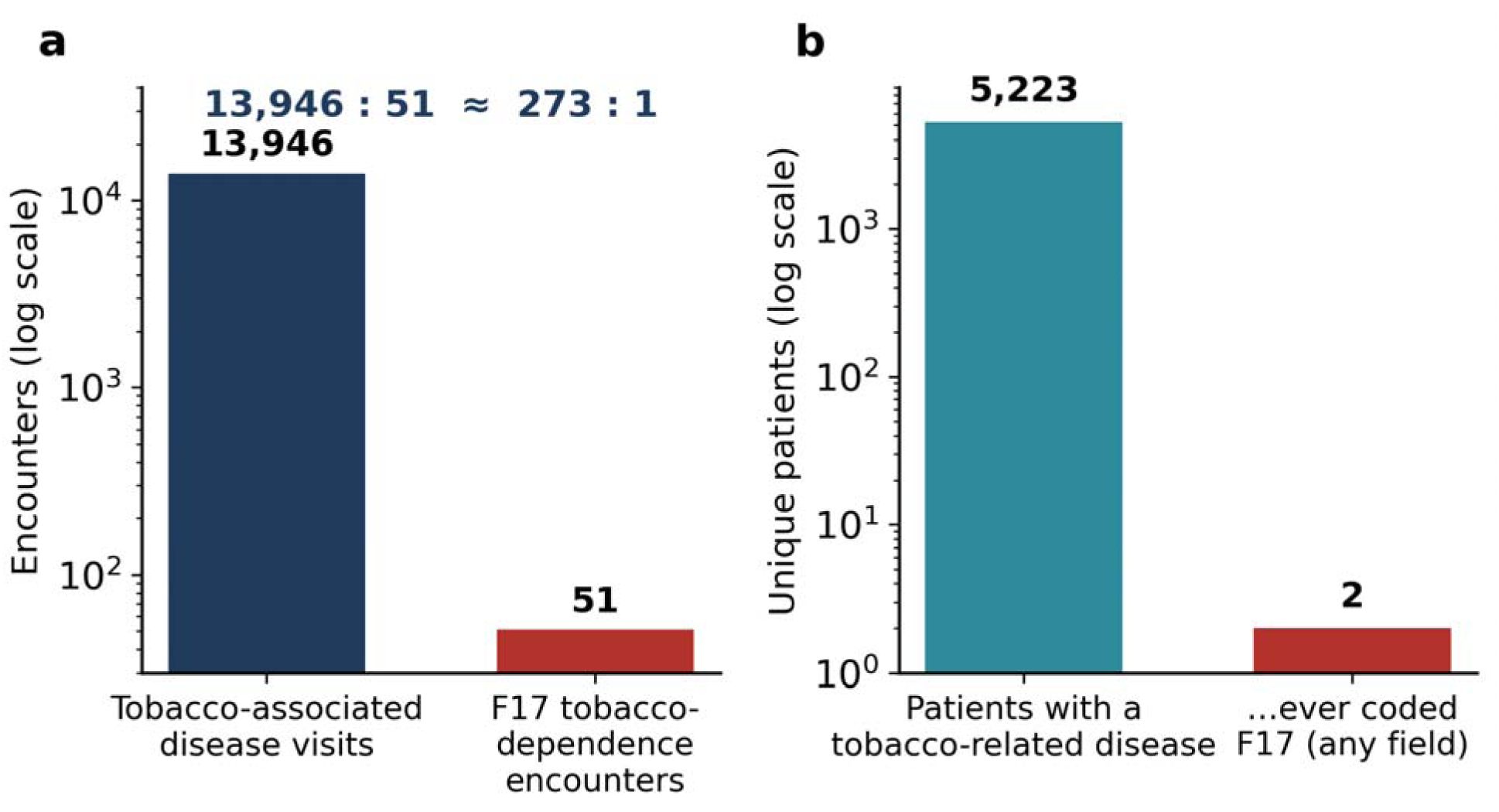
The tobacco cost paradox. Tobacco-associated disease visits (13,946) versus F17 tobacco-dependence encounters (51) on a logarithmic scale (≈273:1), and the number of disease patients ever coded F17 (2 of 5,223).

### Trends over time

The tobacco-associated disease burden rose steeply over the study period (Figure 4): annual disease-group visits increased from 757 in 2015 to 3,979 in 2023, and verified annual costs from US$0.14 million to US$1.63 million. F17 coding did not increase commensurately and remained negligible throughout, so the disease-to-F17 gap widened rather than narrowed over the nine years.

**Figure 4.**
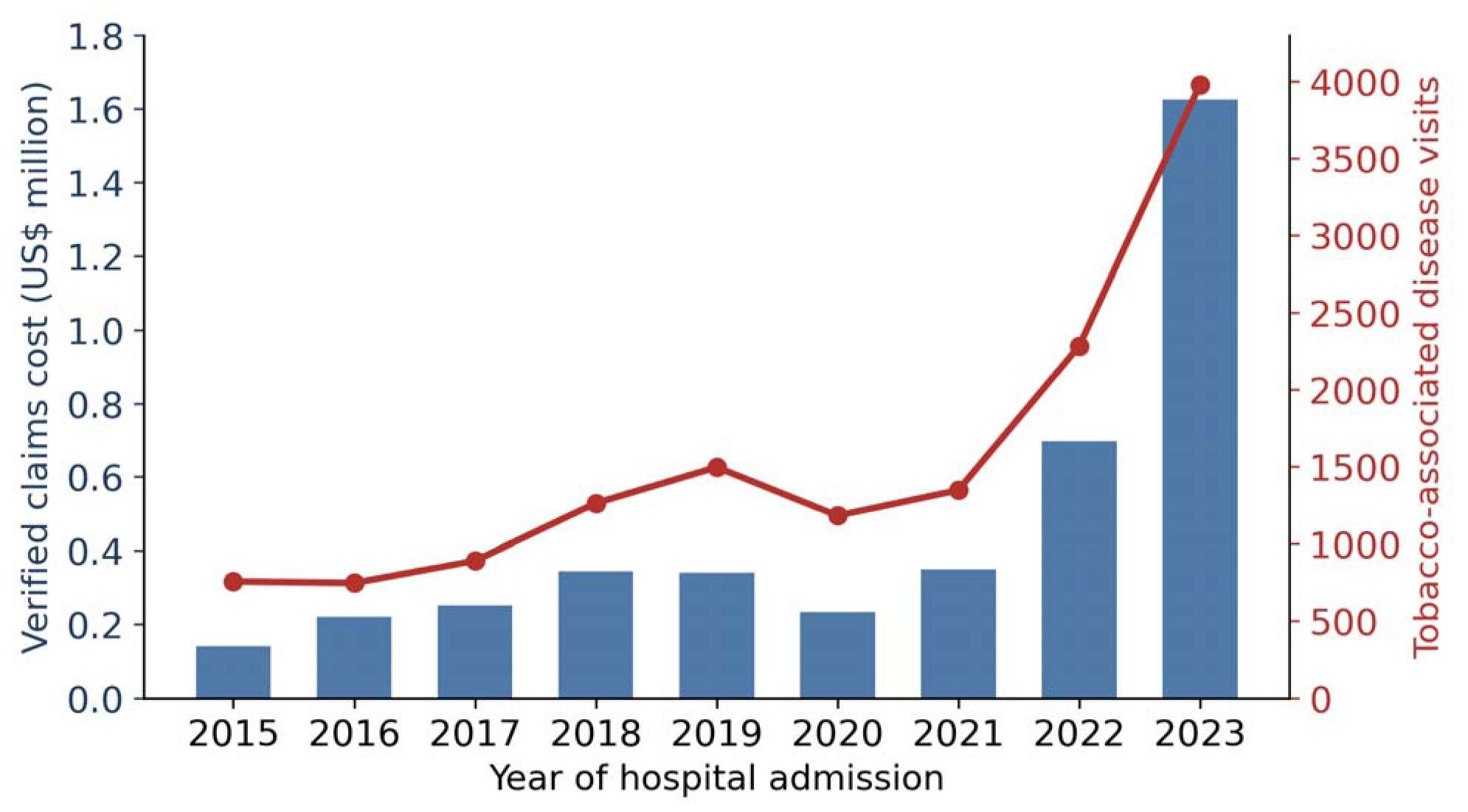
Rising tobacco-associated disease burden in the national mental-health sample, 2015–2023: annual verified cost in US$ million (bars) and disease-group visits (line). Note for submission: per BMJ Open guidance, figures are provided as separate high-resolution image files (≥300 dpi) and uploaded separately rather than embedded. They are reproduced below for reviewer convenience only.

### Sensitivity analyses

The central contrast was robust to the pre-specified alternative case definitions. Ascertaining tobacco-associated disease from any hospital diagnosis field rather than the primary field alone increased disease visits from 13,946 to 117,690 and verified cost from US$4.20 to 6.85 million, widening the disease-to-F17 ratio from approximately 273:1 to 2,308:1; even under this broadest definition only 4 of 8,519 disease patients (0.05%) were ever assigned an F17 code. Broadening COPD from J44 to J40–J44 changed the totals only marginally (15,300 visits; US$4.33 million; ratio approximately 300:1), and excluding claims with missing or non-positive verified cost left the base-case results unchanged. The paradox therefore does not depend on the particular ICD-10 grouping or on the use of the primary-diagnosis field; the absolute figures are definition-dependent, but the direction and order of magnitude are not.

## Discussion

### Key results

In a national health-insurance sample of more than one million weighted members with mental and behavioural disorders, the insurer paid for a large and rising volume of tobacco-associated hospital care—13,946 verified visits and US$4.20 million (weighted ∼US$74.7 million)—while the underlying tobacco dependence was almost never recorded. With 51 hospital F17 encounters against 13,946 disease visits (≈273:1), and only 2 of 5,223 disease patients ever coded F17, the insurer is, in effect, financing the consequences of tobacco use without registering the addiction that drives them. We term this the tobacco cost paradox: visible, expensive disease and invisible, untreated dependence.

### Comparison with other studies

The dominance of cardiovascular and cerebrovascular disease in our cost data (95% of the four-group total) is consistent with national-claims and hospital costing studies elsewhere: cardiometabolic disease accounted for 53% of smoking-attributable spending in South Korea’s NHIS^9^ and cardiovascular disease for roughly three-quarters of smoking-attributable hospital costs in Iran.^11^ Our finding of near-absent dependence coding echoes evidence that administrative systems systematically under-record tobacco: United States Medicare tobacco-use coding remained well below survey prevalence,^5^ and Medicaid analyses document low tobacco-use-disorder diagnosis and cessation-treatment billing,^6^ including among the substance-use and mental-health populations who smoke most and benefit most from coverage.^7^ Our results extend this literature to a large Southeast Asian single-payer system and, unusually, place the cost of disease and the coding of dependence side by side in the same population.

### Interpretation and the use of routinely collected data

Two interpretations of the paradox are possible, and both carry the same policy implication. Either tobacco dependence is genuinely under-treated in this population, or it is treated informally but not coded or reimbursed; in either case, dependence is not being captured as a financeable clinical activity. Because F17 carries no reimbursement advantage and cessation pharmacotherapy is not covered, clinicians have little incentive to code it—a classic limitation of routinely collected data, in which absence of a code reflects the incentives of the payment system as much as the patient’s clinical state. The corollary is encouraging: the same claims architecture that today renders addiction invisible could render it visible and actionable if F17 coding, brief cessation intervention and reimbursable pharmacotherapy were built into primary-care pathways.

### Policy context: a reimbursement gap, not a smoker exclusion

Our findings should not be read as evidence that the national insurance scheme penalises smokers. The scheme does not exclude smokers or smoking-related illness—its administrator has publicly stated that no regulation withholds cover from people who smoke—and our data show precisely that it pays, heavily, for smoking-associated COPD, cancer, ischaemic heart disease and stroke. What the benefit package does not finance is treatment of the dependence that drives those diseases. Under the Presidential Regulation governing the scheme (Perpres 82/2018, amended by Perpres 59/2024), the services explicitly excluded from cover include disorders arising from drug and alcohol dependency; tobacco dependence is not itself listed, but neither its treatment nor cessation care is included as a reimbursable benefit.^18,19^ Cessation pharmacotherapy—nicotine-replacement therapy, bupropion and varenicline—is not covered, and cessation support is provided only as promotive or preventive counselling (the Ministry of Health “Upaya Berhenti Merokok” programme and a national quit-line), outside the curative, case-based payment stream.^20^ Because a standalone F17 diagnosis attracts no meaningful tariff, clinicians have neither a clear pathway nor a financial incentive to record or treat dependence—exactly the pattern of near-absent F17 coding we observed.

The population this gap fails most is the one studied here. People with severe mental illness, who dominate this cohort, smoke at two to three times the rate of the general population and suffer a disproportionate share of tobacco-related death, yet are least able to fund cessation care themselves.^8^ The system meets them at the costly downstream end—stroke units, cardiac wards and oncology services—while offering nothing reimbursable at the upstream, treatable stage of dependence, perpetuating the very morbidity it later pays to treat.

Closing this gap is feasible and, set against the costs documented here, plausibly cost-saving. Concrete steps include adding brief cessation counselling and reimbursable cessation pharmacotherapy to the primary-care benefit package; attaching a payable tariff or capitation incentive to F17 recording so that dependence becomes a financed clinical activity; embedding routine smoking-status screening and cessation referral into the chronic-disease and mental-health pathways that already see these patients; and earmarking part of tobacco-excise revenue to fund cessation services. A modest upstream investment could be offset against the large and rising downstream expenditure shown in this study.

### Limitations

Several limitations follow from the data. First, administrative claims cannot establish that any individual case was caused by smoking; we report the cost of tobacco-associated diseases, not a smoking-attributable fraction, and we deliberately avoided causal attribution. Second, the dataset contains no smoking status, pack-years or behavioural information, so true tobacco-dependence prevalence cannot be measured—our F17 counts describe coding and reimbursement practice, not epidemiology. Third, this is a mental-health (psychiatric) sample.

People with severe mental illness die younger, are subject to diagnostic overshadowing and have poorer access to specialist cardiac and oncology care, so the severity and cost of tobacco-associated disease captured here may understate the true burden in this group; conversely, restricting the sample to mental-health service users means the estimates cannot be extrapolated to the general smoking population, in whom the absolute burden is far larger.

Fourth, choices of ICD-10 grouping influence absolute counts; we pre-specified and published the code list and tested alternative definitions in sensitivity analyses, which left the direction and order of magnitude of the contrast unchanged. Fifth, the data were not created to answer our question, and enrolment, case ascertainment and coding completeness expanded substantially over 2015–2023 as the scheme matured; part of the steep rise in tobacco-associated disease cost therefore reflects growing coverage and more complete coding rather than a true increase in incidence. Finally, all costs were converted at a single period-average exchange rate; year-specific or purchasing-power-parity conversion would shift the absolute US-dollar figures, though not the disease-to-F17 contrast, and weighted projections depend on the sampling weights and should be read as population estimates rather than exact totals.

### Generalisability

Within Indonesia, the findings are most directly applicable to insured members with mental and behavioural disorders—a group with high smoking prevalence and high downstream cost—but the structural driver, the absence of a reimbursement pathway for tobacco dependence, is a feature of the national scheme rather than of this subsample, and is therefore likely to operate system-wide. The pattern is consistent with under-coding observed in other single-payer and public-insurance systems internationally.^5,6,9^

## Conclusion

In Indonesia’s national health insurance scheme, tobacco-associated disease is an expensive and growing line item while tobacco dependence is almost never coded or treated. Integrating routine F17 coding, brief cessation consultation and reimbursable cessation pharmacotherapy into primary care would make addiction visible within the claims system and could convert a large downstream cost centre into an upstream addiction-care target. Future work linking smoking-attributable fractions to claims-based cost data, and evaluating the budget impact of covering cessation treatment, would help quantify the return on such an investment.

## Statements

### Ethics approval

This study analysed a de-identified, governance-controlled secondary dataset released by BPJS Kesehatan and did not involve identifiable individuals; individual patient consent was therefore not required. Use of the data was subject to BPJS Kesehatan data-use governance.

### Patient consent for publication

Not applicable; the manuscript contains no identifiable individual data.

## Data Availability

All data produced in the present study are available upon reasonable request to the authors

## Author contributions

AN and AJ conceived and designed the study. AJ curated the data and conducted the statistical analysis. AN and AJ interpreted the results. AN drafted the manuscript; AJ critically revised it for important intellectual content. Both authors approved the final version and are accountable for the accuracy and integrity of the work.

## Data availability statement

Individual-level claims data are not publicly available because access is restricted by BPJS Kesehatan data-use governance, which requires registration and a data-use agreement with the data custodian; no research-ethics-committee approval was required for this secondary analysis of a de-identified dataset. The ICD-10 code list, claims-selection flow, data-cleaning rules, weighting approach and analysis code are provided as supplementary materials to support reproducibility, subject to these data-governance restrictions.

## Acknowledgements

The authors thank BPJS Kesehatan for access to the de-identified Mental Health Contextual Sample.

Supplementary Figure S1. Data linkage and claims-selection flow, national mental-health claims sample 2015–2023. Five source files (membership, referral-hospital claims, hospital secondary diagnoses, and primary-care capitation and non-capitation) were linked deterministically at member level and encounter level, with 100% record matching across all files and exact agreement of claim and membership sampling weights. The sample of 54,820 members (weighted 1,032,022) generated 2,074,277 referral-hospital visits, of which 51,026 members (93.1%) had at least one hospital claim; a primary diagnosis was missing or invalid in 47 visits (0.002%). Primary-diagnosis screening identified four tobacco-associated disease groups (13,946 visits; 5,223 patients; US$4.20 million verified), contrasted with ICD-10 F17 coding (51 hospital encounters; 15 patients), giving a disease-to-F17 ratio of approximately 273:1. Sensitivity analyses under alternative case definitions are also summarised. Provided as a separate high-resolution image file (Supplementary_Figure_S1_linkage_flow.png).

