## Supplementary Materials for "Tobacco-Associated Disease Claims and ICD-10 F17 Tobacco-Dependence Coding Among Psychiatric Patients in Indonesia’s National Health Insurance Dataset: A Retrospective Claims-Based Observational Study, 2015–2023"

These supplementary materials accompany the manuscript above. They provide the ICD-10 code list, the claims-selection flow, the data-cleaning and linkage rules, the weighting and variance-estimation approach, the sensitivity analyses and the complete reproducible analysis code, to support reproducibility of the study — subject to the BPJS Kesehatan data-use governance restrictions set out in Section S9 and in the manuscript’s data-availability statement.

**Currency.** All costs are reported in US dollars (US$), converted from Indonesian rupiah (IDR) at the 2015–2023 average official exchange rate of IDR 14,200 per US$1 (World Bank). A single period-average rate was applied to all years so that reported changes reflect health-care activity rather than exchange-rate movement.

**Contents:** S1 ICD-10 code list · S2 Data files and variable dictionary · S3 Claims-selection flow · S4 Data-cleaning and linkage rules · S5 Missing and invalid data · S6 Weighting and variance estimation · S7 Sensitivity analyses · S8 Trends over time · S9 Data-governance statement · S10 Reproducible analysis code · Figure S1.

### S1. ICD-10 code list for case identification

Tobacco-associated disease groups were defined a priori from the referral-hospital primary-diagnosis field (FKL17A; three-character ICD-10 code). Tobacco dependence (F17) was ascertained from the hospital primary (FKL17A) and secondary (FKL24A) diagnosis fields and from the primary-care diagnosis fields (capitation FKP14A/FKP15; non-capitation PNK13A/PNK14). Codes were used as recorded by providers and were not independently validated against medical records.

**Table S1.** ICD-10 code list and diagnosis fields searched for case and exposure identification.

| **Condition group** | **ICD-10 code(s)** | **Diagnosis field(s) searched** | **Source file (variable)** |
| --- | --- | --- | --- |
| Chronic obstructive pulmonary disease (COPD) | J44 (base); J40–J44 (sensitivity) | Hospital primary diagnosis | fkrtl (FKL17A) |
| Trachea, bronchus and lung cancer | C33, C34 | Hospital primary diagnosis | fkrtl (FKL17A) |
| Ischaemic heart disease | I20–I25 | Hospital primary diagnosis | fkrtl (FKL17A) |
| Stroke / cerebrovascular disease | I60–I69 | Hospital primary diagnosis | fkrtl (FKL17A) |
| Tobacco dependence (exposure of interest) | F17 | Hospital primary & secondary; primary-care diagnosis | fkrtl (FKL17A), fkrtldxsekunder (FKL24A), fktpkapitasi (FKP14A/FKP15), fktpnonkapitasi (PNK13A/PNK14) |
| Mental/behavioural disorders (cohort definition) | F00–F99 | All hospital & primary-care diagnosis fields | all diagnosis fields above |

***Note:*** *Codes were matched at the three-character level for grouping; numeric ICD-10 ranges (I20–I25, I60–I69, J40–J44) were parsed from the letter–number prefix. The qualitative disease-to-F17 contrast is robust to reasonable alternative groupings (Section S7).*

### S2. Source data files and variable dictionary

The analysis used the complete released Mental Health Contextual Sample (Sampel Kontekstual Kesehatan Mental, BPJS Kesehatan) — five linked files. Files were linked at member level by the anonymised member identifier (PSTV01) and at encounter level by the hospital visit identifier (FKL02).

**Table S2.** Source data files and record counts.

| **File** | **Contents** | **Records** |
| --- | --- | --- |
| KM2023_kepesertaan.dta | Membership: demographics, sampling weight, vital status | 54,820 members |
| KM20152023_fkrtl.dta | Referral-hospital (FKRTL) claims: primary diagnosis, service date, verified cost | 2,074,277 |
| KM20152023_fkrtldxsekunder.dta | Hospital secondary diagnoses (linked to visit by FKL02) | 2,240,698 |
| KM20152023_fktpkapitasi.dta | Primary-care capitation claims | 1,743,537 |
| KM20152023_fktpnonkapitasi.dta | Primary-care non-capitation claims | 2,486 |

**Table S3.** Variable dictionary for fields used in the analysis.

| **Variable** | **Label / role** | **File(s)** |
| --- | --- | --- |
| PSTV01 | Anonymised member identifier (member-level linkage key) | all files |
| PSTV15 | Member sampling weight (pweight) | all files |
| PSTV10 | District of residence (survey stratum) | kepesertaan |
| PSTV05 | Sex | kepesertaan |
| PSTV03 | Date of birth (age computed at 2023) | kepesertaan |
| PSTV07 | Hospital ward-class entitlement | kepesertaan |
| PSTV08 | Contribution segment (PBI / non-PBI) | kepesertaan |
| PSTV18 | Vital status / year of death | kepesertaan |
| FKL02 | Hospital visit identifier (encounter-level linkage key) | fkrtl, fkrtldxsekunder |
| FKL03 | Service date | fkrtl |
| FKL17A | Hospital primary diagnosis (3-character ICD-10) | fkrtl |
| FKL24A | Hospital secondary diagnosis (ICD-10) | fkrtldxsekunder |
| FKL47 | Provider-billed amount (not used) | fkrtl |
| FKL48 | Insurer-verified payable amount (cost outcome) | fkrtl |
| FKP14A / FKP15 | Primary-care (capitation) diagnosis fields | fktpkapitasi |
| PNK13A / PNK14 | Primary-care (non-capitation) diagnosis fields | fktpnonkapitasi |

### S3. Claims-selection flow

The data-linkage and claims-selection process, with record counts and linkage quality at each stage, is shown in Figure S1 (reproduced at the end of this document) and summarised in Figure 1 of the manuscript.

**Stage 1 — Source population.** 54,820 members of the JKN Mental Health Contextual Sample (weighted 1,032,022), each with ≥1 recorded ICD-10 F-code (F00–F99) diagnosis and a BPJS sampling weight (PSTV15).

**Stage 2 — Linked hospital claims.** 2,074,277 referral-hospital (FKRTL) visits (weighted 40,196,048; total verified cost US$70.6 million), linked to members by PSTV01 and to secondary diagnoses by visit ID FKL02. Of the 54,820 members, 51,026 (93.1%) had ≥1 hospital claim.

**Stage 3 — Disease ascertainment.** Visits with a primary diagnosis in one of the four tobacco-associated disease groups → 13,946 visits among 5,223 patients (US$4.20 million verified; ~6.0% of the sample’s hospital spending).

**Stage 4 — Exposure ascertainment.** F17 tobacco-dependence codes searched in hospital primary (n=4) and secondary (n=47) fields (51 encounters, 15 patients) and in primary care (26 capitation visits, 17 patients; 0 non-capitation) — 77 F17-coded encounters in total across all settings over nine years.

**Contrast.** 13,946 disease visits vs 51 hospital F17 encounters ≈ 273:1; only 2 of 5,223 disease patients (0.04%) were ever assigned an F17 code in any field or setting.

### S4. Data-cleaning and linkage rules

- One record per verified hospital visit (unique FKL02); distinct visits were not de-duplicated.
- Cost defined as the BPJS-verified payable amount (FKL48), used in preference to the provider-billed amount (FKL47).
- Disease grouping restricted to the primary-diagnosis field to limit rule-out and historical (e.g. Z-code) misclassification; records with a missing/invalid primary diagnosis were retained in denominators but assigned to no disease group.
- ICD-10 codes were trimmed and upper-cased; three-character grouping applied; numeric ICD-10 ranges parsed from the letter–number prefix.
- Service dates parsed to calendar year; age computed at 2023 from date of birth (used for age summaries only).
- No records were imputed or deleted during cleaning; linkage was deterministic (member level on PSTV01; secondary-diagnosis file on FKL02).

**Table S4.** Linkage quality across the five source files (RECORD 12.3).

| **Linked file** | **Records** | **Matched** | **%** |
| --- | --- | --- | --- |
| Referral-hospital (fkrtl) → membership (PSTV01) | 2,074,277 | 2,074,277 | 100% |
| Primary-care capitation → membership | 1,743,537 | 1,743,537 | 100% |
| Primary-care non-capitation → membership | 2,486 | 2,486 | 100% |
| Hospital secondary dx → visit (FKL02) | 2,240,698 | 2,240,698 | 100% |

***Note:*** *Claim-level and membership sampling weights agreed exactly for every record. 51,026 of 54,820 members (93.1%) had at least one referral-hospital claim during 2015–2023.*

### S5. Missing and invalid data

**Table S5.** Missing or invalid values in fields used for description and analysis (STROBE 14b, RECORD 13.1).

| **Field** | **Missing / invalid** | **%** |
| --- | --- | --- |
| Membership: sex, date of birth, ward class, contribution segment, district, weight | 0 | 0.000% |
| Hospital primary diagnosis (FKL17A) | 47 of 2,074,277 | 0.002% |
| Service date (FKL03) | 0 | 0.000% |
| Verified cost (FKL48) — missing | 2 | <0.001% |
| Verified cost (FKL48) — non-positive | 3 | <0.001% |

***Note:*** *Records with a missing/invalid primary diagnosis or a non-positive cost were retained in denominators but could not enter a disease group; none were imputed or deleted. No records were excluded for missing covariates.*

### S6. Weighting and variance estimation

All weighted estimates apply the member sampling weight PSTV15 (mean 18.8; range 0.98–34.82). Weighted member and patient counts were obtained by summing weights over unique members; weighted visit counts by summing weights over records; and weighted costs by summing the product of verified cost (FKL48) and weight. Population-weighted projections should be read as population estimates rather than exact totals.

For member-level proportions, 95% confidence intervals were estimated with a Taylor-linearised survey (design-based) estimator, using the member (PSTV01) as the primary sampling unit and district of residence (PSTV10) as the stratum, matching the insurer’s declared design:

svyset PSTV01 [pweight=PSTV15], strata(PSTV10), vce(linearized), singleunit(certainty)

All reported estimates are descriptive and unadjusted; because the study quantifies coded and reimbursed activity rather than testing an exposure–outcome association, no adjustment for confounders was undertaken. Continuous age is summarised as median and interquartile range.

### S7. Sensitivity analyses

Three sensitivity analyses were pre-specified to test the robustness of the disease-to-F17 contrast to coding choices: (i) ascertaining disease from any hospital diagnosis field (primary or secondary) rather than the primary field alone; (ii) broadening COPD from J44 to J40–J44; and (iii) excluding claims with missing or non-positive verified cost. The hospital F17 denominator is 51 encounters throughout.

**Table S6.** Sensitivity analyses for the disease-to-F17 contrast (US$).

| **Analysis** | **Disease visits** | **Patients** | **Verified cost** | **Disease:F17 ratio** | **Patients ever F17** |
| --- | --- | --- | --- | --- | --- |
| Base case (primary dx; J44 / C33–C34 / I20–I25 / I60–I69) | 13,946 | 5,223 | US$4.20M | ≈273:1 | 2 (0.04%) |
| S1 — any diagnosis field (primary or secondary) | 117,690 | 8,519 | US$6.85M | ≈2,308:1 | 4 (0.05%) |
| S2 — broadened COPD (J40–J44) | 15,300 | — | US$4.33M | ≈300:1 | — |
| S3 — exclude missing/non-positive cost | 13,946 | 5,223 | US$4.20M | ≈273:1 | 2 (0.04%) |

***Note:*** *“Patients ever F17” is the number of disease patients ever assigned an F17 code in any field or setting (percentage of disease patients). The direction and order of magnitude of the contrast are unchanged across all definitions; absolute figures are definition-dependent. Dashes denote values not separately reported for that analysis.*

### S8. Trends over time

The tobacco-associated disease burden rose steeply over the study period while F17 coding remained negligible throughout, so the disease-to-F17 gap widened rather than narrowed. The first- and last-year values are shown below; the full year-by-year series is regenerated by analysis_code.py (Section S10).

**Table S7.** Tobacco-associated disease burden, first vs last study year.

| **Year** | **Disease-group visits** | **Verified cost (US$ million)** |
| --- | --- | --- |
| 2015 | 757 | 0.14 |
| 2023 | 3,979 | 1.63 |

***Note:*** *Part of the rise reflects expanding coverage and more complete coding as the scheme matured over 2015–2023, not necessarily a true increase in incidence (see manuscript Limitations).*

### S9. Data-governance and availability statement

Individual-level JKN claims data are not publicly available because access is restricted by BPJS Kesehatan data-use governance, which requires registration and a data-use agreement with the data custodian. The dataset analysed is a de-identified, governance-controlled sample released by BPJS Kesehatan and contains no directly identifying information; no research-ethics-committee approval was required for this secondary analysis of a de-identified dataset. The ICD-10 code list, claims-selection flow, data-cleaning rules, weighting approach and analysis code are provided in these supplementary materials to support reproducibility within these governance constraints. No attempt was made to re-identify individuals.

### S10. Reproducible analysis code

The complete analysis was conducted in Python 3 (pandas, NumPy; pyreadstat for Stata-format input). The script below regenerates every count, cost and confidence interval reported in the manuscript and figures — the cohort description, disease-group visits/patients/costs, F17 coding counts, the disease-to-F17 ratio, weighted survey-design prevalence with 95% CIs, linkage-quality and missing-data checks, the three sensitivity analyses, and the annual trend — from the source claim files. Set DATA_DIR to the folder containing the KM20152023_* .dta files. The script is also provided as a separate file, analysis_code.py.

```
#!/usr/bin/env python3
# =============================================================================
# Reproducible analysis
# Tobacco-associated disease claims vs ICD-10 F17 tobacco-dependence coding
# JKN Mental Health Contextual Sample (BPJS Kesehatan), 2015-2023
#
# Manuscript: "Tobacco-Associated Disease Claims and ICD-10 F17 Tobacco-
#   Dependence Coding Among Psychiatric Patients in Indonesia's National Health
#   Insurance Dataset: A Retrospective Claims-Based Observational Study,
#   2015-2023."
#
# This single script regenerates every count, cost, confidence interval,
# linkage/missing-data check, sensitivity analysis and annual trend reported in
# the manuscript, figures and supplementary materials.
#
# Costs are reported in US dollars (US$), converted from Indonesian rupiah at the
# 2015-2023 average official exchange rate of IDR 14,200 per US$1 (World Bank).
#
# Requirements: python3, pandas, numpy, pyreadstat
# Usage: set DATA_DIR to the folder containing the KM20152023_* .dta files.
# =============================================================================
import re
import numpy as np
import pandas as pd
import pyreadstat

DATA_DIR = "."          # <-- set to the BPJS data folder
USD = 14200.0           # IDR per US$1 (2015-2023 period-average official rate)

FK  = f"{DATA_DIR}/KM20152023_fkrtl.dta"            # referral-hospital claims
SEC = f"{DATA_DIR}/KM20152023_fkrtldxsekunder.dta"  # hospital secondary diagnoses
MEM = f"{DATA_DIR}/KM2023_kepesertaan.dta"          # membership / weights
KAP = f"{DATA_DIR}/KM20152023_fktpkapitasi.dta"     # primary care (capitation)
NK  = f"{DATA_DIR}/KM20152023_fktpnonkapitasi.dta"  # primary care (non-capitation)


# ---- Helpers ----------------------------------------------------------------
def norm(s):
    """Trim and upper-case an ICD-10 string column; missing -> ''. """
    return s.astype("string").str.strip().str.upper().fillna("")


def disease_group(code, broad=False):
    """Map a three-character ICD-10 code to a tobacco-associated disease group.
    broad=True widens COPD from J44 to J40-J44 (sensitivity analysis 2)."""
    if not isinstance(code, str) or len(code) < 2:
        return None
    if broad:
        m = re.match(r"^J(\d{2})", code)
        if m and 40 <= int(m.group(1)) <= 44:
            return "COPD"
    else:
        if code == "J44":
            return "COPD"
    if code in ("C33", "C34"):
        return "Lung/bronchus/trachea cancer"
    m = re.match(r"^I(\d{2})", code)
    if m:
        n = int(m.group(1))
        if 20 <= n <= 25:
            return "Ischaemic heart disease"
        if 60 <= n <= 69:
            return "Stroke (cerebrovascular)"
    return None


# ---- Load -------------------------------------------------------------------
mem, _ = pyreadstat.read_dta(MEM, usecols=["PSTV01", "PSTV03", "PSTV05", "PSTV07",
                                           "PSTV08", "PSTV10", "PSTV15", "PSTV18"])
fk, _ = pyreadstat.read_dta(FK, usecols=["PSTV01", "PSTV15", "FKL02", "FKL03",
                                         "FKL17A", "FKL47", "FKL48"])
sec, _ = pyreadstat.read_dta(SEC, usecols=["FKL02", "FKL24A"])

fk["c3"] = norm(fk["FKL17A"])
fk["grp"] = fk["c3"].map(lambda x: disease_group(x, broad=False))
fk["grp_broad"] = fk["c3"].map(lambda x: disease_group(x, broad=True))
fk["year"] = pd.to_datetime(fk["FKL03"], errors="coerce").dt.year
sec["s3"] = norm(sec["FKL24A"])

usd = lambda idr: idr / USD          # IDR -> US$
musd = lambda idr: idr / USD / 1e6   # IDR -> US$ million

# ---- Cohort -----------------------------------------------------------------
print("=" * 70); print("COHORT")
print(f"Members              : {len(mem):>12,} | weighted {mem.PSTV15.sum():>14,.0f}")
print(f"Hospital visits      : {len(fk):>12,} | weighted {fk.PSTV15.sum():>14,.0f}")
print(f"Total verified cost  : US$ {usd(fk.FKL48.sum()):>14,.0f}  "
      f"(US$ {musd(fk.FKL48.sum()):.1f}M)")

# ---- Disease groups (base case: primary diagnosis only) ---------------------
print("\n" + "=" * 70); print("DISEASE GROUPS (primary-diagnosis field, US$)")


def w_unique_patients(d):
    return d.drop_duplicates("PSTV01").PSTV15.sum()


groups = ["COPD", "Lung/bronchus/trachea cancer",
          "Ischaemic heart disease", "Stroke (cerebrovascular)"]
rows = []
for g in groups:
    d = fk[fk.grp == g]
    rows.append(dict(group=g, visits=len(d), patients=d.PSTV01.nunique(),
                     cost_usd=usd(d.FKL48.sum()), median_usd=usd(d.FKL48.median()),
                     w_visits=d.PSTV15.sum(), w_patients=w_unique_patients(d),
                     w_cost_usd=usd((d.FKL48 * d.PSTV15).sum())))
allg = fk[fk.grp.notna()]
rows.append(dict(group="ALL FOUR", visits=len(allg), patients=allg.PSTV01.nunique(),
                 cost_usd=usd(allg.FKL48.sum()), median_usd=np.nan,
                 w_visits=allg.PSTV15.sum(), w_patients=w_unique_patients(allg),
                 w_cost_usd=usd((allg.FKL48 * allg.PSTV15).sum())))
print(pd.DataFrame(rows).to_string(index=False))

cvd = fk[fk.grp.isin(["Ischaemic heart disease", "Stroke (cerebrovascular)"])]
print(f"\nCVD+cerebro verified cost: US$ {usd(cvd.FKL48.sum()):,.0f} "
      f"(US$ {musd(cvd.FKL48.sum()):.2f}M) | weighted US$ "
      f"{musd((cvd.FKL48 * cvd.PSTV15).sum()):.1f}M")
print(f"Share of four-group cost : "
      f"{100 * cvd.FKL48.sum() / allg.FKL48.sum():.0f}%")
print(f"Four-group share of all hospital spend: "
      f"{100 * allg.FKL48.sum() / fk.FKL48.sum():.1f}%")

# ---- F17 tobacco-dependence coding ------------------------------------------
print("\n" + "=" * 70); print("F17 TOBACCO-DEPENDENCE CODING")
f17_primary = fk[fk.c3 == "F17"]
f17_sec_visits = set(sec.loc[sec.s3 == "F17", "FKL02"])
v2p = fk[["FKL02", "PSTV01"]].drop_duplicates()
f17_patients = set(v2p.loc[v2p.FKL02.isin(set(f17_primary.FKL02) | f17_sec_visits),
                           "PSTV01"])
print(f"F17 primary  (hospital): {len(f17_primary)}")
print(f"F17 secondary(hospital): {len(f17_sec_visits)}")
print(f"F17 any hospital       : {len(f17_primary) + len(f17_sec_visits)} encounters "
      f"| {len(f17_patients)} patients")

kap, _ = pyreadstat.read_dta(KAP, usecols=["PSTV01", "PSTV15", "FKP14A", "FKP15"])
nk, _ = pyreadstat.read_dta(NK, usecols=["PSTV01", "PSTV15", "PNK13A", "PNK14"])
kap17 = (norm(kap.FKP14A) == "F17") | norm(kap.FKP15).str.startswith("F17")
nk17 = (norm(nk.PNK13A) == "F17") | norm(nk.PNK14).str.startswith("F17")
print(f"F17 primary care (cap) : {int(kap17.sum())} visits "
      f"| {kap.loc[kap17, 'PSTV01'].nunique()} patients")
print(f"F17 primary care (noncap): {int(nk17.sum())} visits")

# ---- Disease-to-F17 ratio + overlap -----------------------------------------
print("\n" + "=" * 70); print("DISEASE-TO-F17 CONTRAST")
disease_visits = len(allg)
hosp_f17 = len(f17_primary) + len(f17_sec_visits)
print(f"Disease visits {disease_visits:,} vs hospital F17 {hosp_f17} "
      f"= {disease_visits / hosp_f17:.0f} : 1")
disease_patients = set(allg.PSTV01)
print(f"Disease patients ever coded F17: "
      f"{len(disease_patients & f17_patients)} of {len(disease_patients):,}")

# ---- Member-level prevalence with survey (Taylor-linearised) 95% CI ---------
print("\n" + "=" * 70); print("WEIGHTED MEMBER-LEVEL PREVALENCE (survey 95% CI)")


def svy_prop_ci(y, w, stratum):
    """Taylor-linearised survey proportion and 95% CI.
    Reproduces: svyset PSTV01 [pweight=PSTV15], strata(PSTV10)."""
    d = pd.DataFrame({"y": np.asarray(y, float), "w": np.asarray(w, float),
                      "h": np.asarray(stratum)}).dropna()
    Nhat = d.w.sum()
    p = (d.w * d.y).sum() / Nhat
    d["g"] = d.w * ((d.y - p) / Nhat)
    var = 0.0
    for _, dh in d.groupby("h"):
        n = len(dh)
        if n < 2:
            continue  # singleton strata treated as certainty (singleunit(certainty))
        var += n / (n - 1) * ((dh.g - dh.g.mean()) ** 2).sum()
    se = var ** 0.5
    return 100 * p, 100 * (p - 1.96 * se), 100 * (p + 1.96 * se)


mdis = allg.groupby("PSTV01").grp.agg(set)
mem = mem.set_index("PSTV01")
for g in groups:
    mem[g] = mem.index.map(lambda i: int(i in mdis.index and g in mdis[i]))
mem["any4"] = mem[groups].max(axis=1)
mem["male"] = (mem.PSTV05 == 1).astype(int)
mem = mem.reset_index()
for lab, col in [("Male", "male")] + [(g, g) for g in groups] + [("Any of 4", "any4")]:
    est = svy_prop_ci(mem[col].values, mem.PSTV15.values, mem.PSTV10.values)
    print(f"  {lab:<30}: {est[0]:6.2f}%  (95% CI {est[1]:.2f}-{est[2]:.2f})")

# ---- Linkage quality (RECORD 12.3) ------------------------------------------
print("\n" + "=" * 70); print("LINKAGE QUALITY (RECORD 12.3)")
memids = set(mem.PSTV01)


def linkpct(df, name):
    matched = df.PSTV01.isin(memids).sum()
    print(f"  {name:<22}: records={len(df):>10,}  "
          f"matched={matched:>10,} ({100 * matched / len(df):.4f}%)")


linkpct(fk, "fkrtl (hospital)")
linkpct(kap, "fktp capitation")
linkpct(nk, "fktp non-capitation")
fkvis = set(fk.FKL02)
secmatch = sec.FKL02.isin(fkvis).sum()
print(f"  {'secondary dx':<22}: records={len(sec):>10,}  "
      f"matched={secmatch:>10,} ({100 * secmatch / len(sec):.4f}%)")
wm = mem.set_index("PSTV01").PSTV15
fkw = fk.dropna(subset=["PSTV01"]).copy()
fkw["memw"] = fkw.PSTV01.map(wm)
agree = np.isclose(fkw.PSTV15, fkw.memw, rtol=1e-4, equal_nan=False).mean()
print(f"  claim weight == membership weight: {100 * agree:.3f}%")
print(f"  members with >=1 hospital claim  : {fk.PSTV01.nunique():,} of {len(mem):,} "
      f"({100 * fk.PSTV01.nunique() / len(mem):.1f}%)")

# ---- Missing / invalid data (STROBE 14b, RECORD 13.1) -----------------------
print("\n" + "=" * 70); print("MISSING / INVALID DATA")
for c, lab in [("PSTV03", "birth date/age"), ("PSTV05", "sex"),
               ("PSTV07", "ward class"), ("PSTV08", "contribution segment"),
               ("PSTV10", "district/stratum"), ("PSTV15", "weight")]:
    print(f"  {lab:<22}: missing {mem[c].isna().sum():>6,} "
          f"({100 * mem[c].isna().mean():.3f}%)")
blank = (fk.c3 == "").sum()
valid_icd = lambda x: bool(re.match(r"^[A-Z]\d", x)) if isinstance(x, str) else False
invalid = (~fk.c3.map(valid_icd)).sum()
print(f"  primary dx missing/invalid: {invalid:>10,} "
      f"({100 * invalid / len(fk):.4f}%)")
print(f"  service date unparseable  : {fk.year.isna().sum():>10,}")
print(f"  verified cost NaN         : {fk.FKL48.isna().sum():>10,}")
print(f"  verified cost <= 0        : {(fk.FKL48 <= 0).sum():>10,}")

# ---- Sensitivity analyses (STROBE 12e/17) -----------------------------------
print("\n" + "=" * 70); print("SENSITIVITY ANALYSES (US$)")
sec["sgrp"] = sec.s3.map(lambda x: disease_group(x, broad=False))
sec_dis_vis = set(sec.loc[sec.sgrp.notna(), "FKL02"])
anyfield = fk.grp.notna() | fk.FKL02.isin(sec_dis_vis)


def scen(mask, label):
    d = fk[mask]
    v, pat, c = len(d), d.PSTV01.nunique(), usd(d.FKL48.sum())
    ratio = v / hosp_f17 if hosp_f17 else np.nan
    everf17 = len(set(d.PSTV01) & f17_patients)
    print(f"  {label:<42} visits={v:>7,}  patients={pat:>6,}  "
          f"costUS$={c / 1e6:>6.2f}M  ratio={ratio:>6.0f}:1  everF17={everf17}")


scen(fk.grp.notna(), "BASE: primary-dx only")
scen(anyfield, "SENS1: primary OR secondary dx")
scen(fk.grp_broad.notna(), "SENS2: broadened COPD J40-J44")
scen(fk.grp.notna() & (fk.FKL48 > 0), "SENS3: base, cost>0 only")

# ---- Annual trends ----------------------------------------------------------
print("\n" + "=" * 70); print("ANNUAL TRENDS (disease groups, US$)")
ty = allg.groupby("year").agg(visits=("FKL02", "size"), cost=("FKL48", "sum"))
ty["cost_US$_million"] = ty["cost"].map(musd)
print(ty[["visits", "cost_US$_million"]].to_string())
print("\nDONE")
```

### Figure S1. Data linkage and claims-selection flow

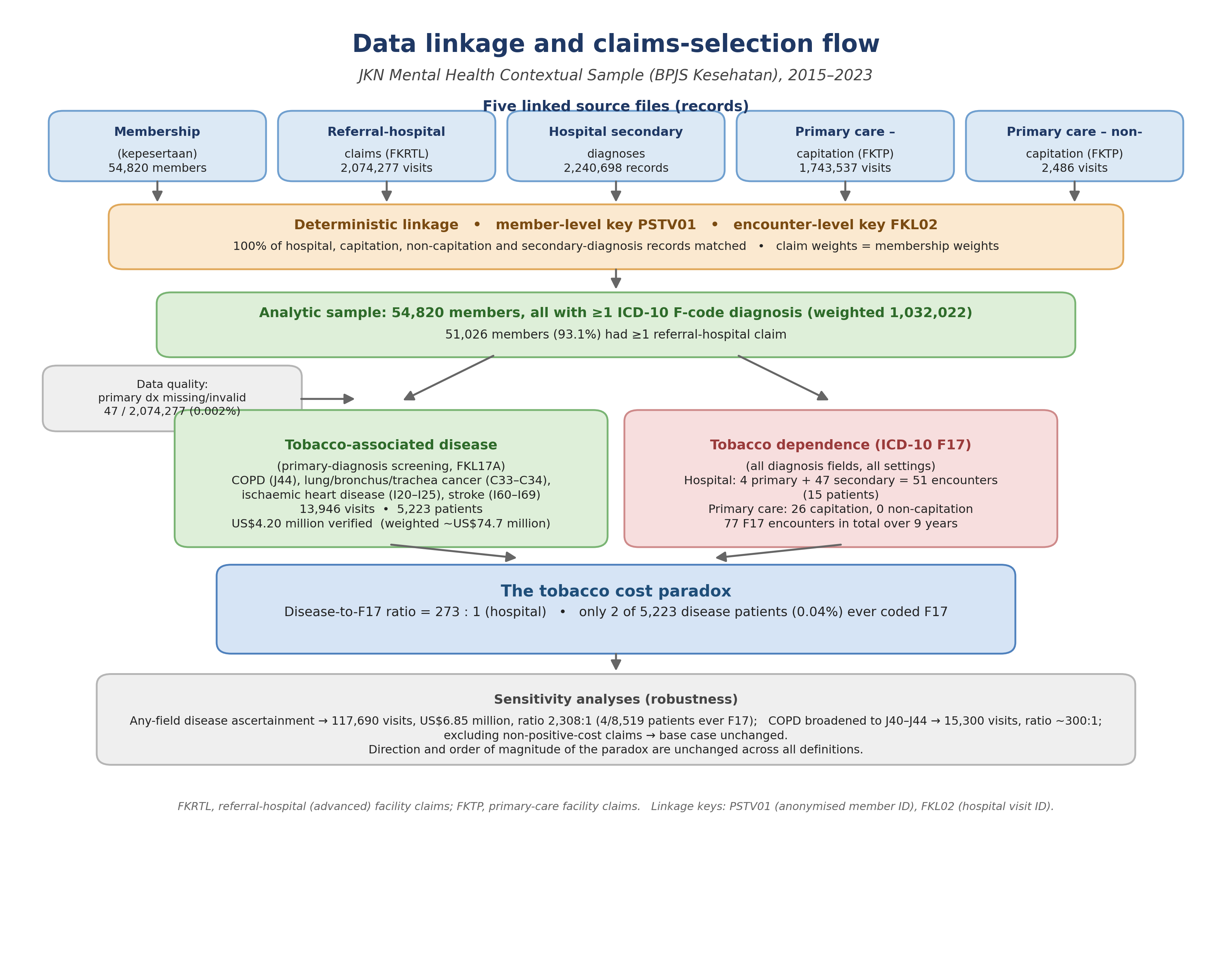

**Figure S1.** Data linkage and claims-selection flow, national mental-health claims sample 2015–2023. Five source files (membership, referral-hospital claims, hospital secondary diagnoses, and primary-care capitation and non-capitation) were linked deterministically at member level (PSTV01) and encounter level (FKL02), with 100% record matching across all files and exact agreement of claim and membership sampling weights. The sample of 54,820 members (weighted 1,032,022) generated 2,074,277 referral-hospital visits, of which 51,026 members (93.1%) had at least one hospital claim; a primary diagnosis was missing or invalid in 47 visits (0.002%). Primary-diagnosis screening identified four tobacco-associated disease groups (13,946 visits; 5,223 patients; US$4.20 million verified), contrasted with ICD-10 F17 coding (51 hospital encounters; 15 patients), giving a disease-to-F17 ratio of approximately 273:1. Sensitivity analyses under alternative case definitions are summarised in Table S6. Provided as a separate high-resolution image file (Supplementary_Figure_S1_linkage_flow.png).
